# Vulnerability of Populations to Malaria after Indoor Residual Spraying is Withdrawn from Areas where its Use has Previously Been Sustained. Protocol for a Systematic Review

**DOI:** 10.1101/2022.05.24.22275507

**Authors:** Philip Orishaba, Edward Kayongo, Pastan Lusiba, Caroline Nakalema, Peter Kasadha, Perez Kiirya, Ismael Kawooya, Rhona Mijumbi-Deve

## Abstract

**Background:** With its proven effectiveness, indoor residual spraying (IRS) as a malaria vector control strategy form one of the reliable vector control strategies, especially when at least 80% of the population is covered. However, to date, there is uncertainty regarding the consequences of IRS withdrawal on malaria control when there’s no clear exit strategy in place. Therefore, there is need to comprehensively update literature regarding malaria burden indicators when IRS is withdrawn following sustained use.

**Methods:** This protocol follows the PRISMA guidelines. We performed a systematic search of studies published between 2000 and 2022 in CINAHL, Embase, MEDLINE, ProQuest, PsychInfo, Scopus and OpenGrey. Using pre-set eligibility criteria, studies will be used to identify the relevant ones by two independent reviewers. Title/ abstracts will first be screened and potentially eligible ones screened using their full-text publications. Any conflicts/ discrepancies at the two stages will be resolved through regular discussion sessions. Included studies will be extracted to capture study & patient characteristics and relevant outcomes (malaria incidence and malaria vector abundance). Relevant tools will be used to assess the risk of bias in the studies measuring the impact of withdrawal. A meta-analysis will be performed, if sufficient homogeneity exists; otherwise, data arising will be presented using tables and by employing narrative synthesis techniques. Heterogeneity will be assessed using a combination of visual inspection of the forest plot along with consideration of the chi-square test and I-square statistic results.

**Ethics and dissemination:** Ethics approval is not applicable for this study since no original data will be collected. The results will be disseminated through peer-reviewed publication and conference presentations. Furthermore, this systematic review will inform the design of exit strategies for IRS-based programs in malaria endemic areas.

**PROSPERO registration number:** CRD42022310655

**Strengths and limitations of this study:** To the strength of this study, we hope to produce the first systematic review of the impact of IRS withdrawal comprehensively. Such comprehension will be defined by including studies from different settings, among different age groups and studies assessing malaria burden indicators either among human populations or mosquito populations or both. This review also utilizes systematic and transparent rigorous procedures guided by the Cochrane handbook and will report results as stated by Preferred Reporting Items for Systematic Reviews and Meta-Analyses statement. Furthermore, an experienced information specialist designed the search strategy and customized it to the different data bases that we intend to search. We anticipate some anticipations. In this systematic review, the certainty of the evidence could be limited by few numbers of publications and low quality of studies.

## INTRODUCTION

Until today, a child dies from malaria every 2 minutes globally [1]. Malaria is the leading cause of morbidity and mortality in Uganda, accounting for 30-50% of outpatient visits and 15-20% of hospital admissions with more than 10,500 annual deaths [2]. Furthermore, it is estimated that 11% of children under 12 months lose their lives to malaria every year [3]. The Malaria Control Program in Uganda combines several strategies, such as Indoor Residual Spraying (IRS), long-lasting insecticide-treated nets (LLINs), and the test and treat strategy [3]. Vulnerability to malaria infection has reduced over the years mainly due to these combined strategies, over which IRS is part given its role in diminishing vector (mosquito) populations [4]. The updated Uganda Malaria Reduction and Elimination Strategic Plan of 2021-2025 aims to reduce malaria infections by 50%, morbidity by 50%, and mortality by 75% in 2025. IRS thus forms one of the key strategies available for mosquito control as the globe looks towards malaria elimination/ eradication.

IRS first proved to be effective for malaria vector control when it was implemented using dichlorodiphenyltrichloroethane (DDT) during the global malaria eradication campaign conducted from 1955 to 1969 whereby the population at risk for malaria infection was reduced by 700 million and malaria was eliminated from 37 countries [5, 6]. Having proven to be effective, IRS use was rolled out in Africa too, and the campaigns demonstrating a success across a wide diversity of settings [7-10]. IRS therefore forms one of the most reliable vector control strategies in malaria control. IRS is a highly effective vector control intervention, especially when at least 80% of the population is covered [11].

IRS was reintroduced in 2006 after nearly a period of 40 years. The IRS program started with ten districts in Northern Uganda known to have the highest burden but later shifted to another 14 districts in May 2014 [12]. Indeed, there was a substantial reduction in the malaria burden between 2007 and early 2014 [13, 14]. Although the universal LLIN distribution campaign continued in the first ten districts, malaria epidemics were reported following IRS withdrawal, prompting a single IRS round in response to the upsurge [15, 16]. However, it is not certain whether the malaria case upsurge was due to other causes or a consequence of IRS withdrawal. For example, first, the Uganda IRS program can only cover 10% of the total population due to limited financial resources [12]. Second, over the years, there’s been a shift in insecticide use informed by the reported possibility of resistance to a particular class.

Indeed, not much is known about what happens when a large-scale deployment of IRS in a given area is halted even when insecticide resistance is not an issue of concern. Several formulations of insecticides for IRS have been made and approved by the World Health Organization (WHO), and all these are results of the classes of insecticides i.e., pyrethroids, carbamates, organophosphates, organochlorines and neonicotinoids [17].

For areas that have recorded pyrethroid resistance, non-pyrethroids have been taken up to protect the benefits/ effectiveness of IRS. For instance, in Northern Uganda, pyrethroids such as lambda-cyhalothrin and organochlorides such as dichlorodiphenyltrichloroethane (DDT) were used for the first time in 2007. However, due to reports of pyrethroids resistance, policy makers shifted to carbamates and/ or organophosphates [18]. The first case of malaria resurgence was recorded in some areas in 1960 following discontinuation of the global malaria eradication campaigns, whereby some resurgence case scenarios elicited terrific epidemics that called for repeated rounds of IRS [10]. Northern Uganda in particular recorded a malaria epidemic and case upsurges following withdrawal of IRS from malaria endemic districts [19, 20].

No systematic reviews or meta-analyses have aimed to summarize the evidence related to withdrawing IRS from malaria endemic areas. There is need to provide a comprehensive synthesis of the literature to enhance the current knowledge on malaria control indices related to IRS withdrawal as relevant studies have been conducted in the recent years. Our aim is therefore to conduct a systematic review to identify evidence to demonstrate the impact and challenges in malaria control, that arise when IRS is withdrawn from malaria endemic areas following sustained use.

## METHODS

This systematic review will be conducted and reported following the Preferred Reporting Items for Systematic Reviews and Meta-Analyses (PRISMA) guidelines.

### Research questions

a. What is the impact of withdrawing IRS on malaria control from areas of high malaria transmission intensities following sustained use?
b. What is the duration of protection of populations after IRS is withdrawn following sustained use?

### Eligibility criteria

#### Type of studies

Studies will be included if they include a follow up aspect. As such, we shall exclude case series, cross sectional surveys, case reports and case control studies. There will be no restrictions on data. Researchers may have used primary data, secondary data or administrative data. Studies that assess the residual efficacy of the insecticides may as well be included in order to establish the duration of protection of populations as well as measure the duration after which mosquito populations increase following spraying.

#### Populations

We seek to include studies conducted in areas known to be endemic to malaria as well as those in areas with high malaria transmission intensities and where indoor residual spraying has previously been sustained. The review will include both studies assessing malaria burden indicators either in human populations or mosquito populations or both. There will be no restrictions on the age or group of the human populations. As such, studies conducted among children under five years of age, school age children, general population and pregnant women will all be included. We will include studies from all settings, for example, community and health facilities.

#### Type of exposure

Eligible studies will be those assessing malaria burden indicators following withdraw of indoor residual spraying after any number of consistent spraying rounds which may be 4 – 6 months apart. There are no restrictions on the reasons for withdraw. Reasons may either be related to cost or achieving some level of malaria vector control.

#### Comparisons

All eligible studies will be required to have a comparison. The comparison will be estimates of malaria indicators obtained when IRS is being implemented consistently or shortly after the implementation. Such a control will be labelled as the baseline whereas the end-line will be estimates obtained to represent the malaria burden when IRS will have been withdrawn. If with a justifiable reason, studies may also be included if the comparison are outcome indicators measured before IRS is ever implemented in an area.

#### Outcome types

Among human populations, the outcomes will be the malaria burden. Malaria burden indicators will be malaria incidence i.e., incidence rates, relative risks, slide positivity rates, entomological inoculation rates, test positivity rates. Among the mosquito populations, the outcome will be malaria vector abundance. Malaria vector abundance indicators will be sporozoite rates and mosquito mortality rates. Studies will be included if they provide at least reasonable/ sufficient data on the primary outcomes.

#### Information sources

A systematic search was performed in CINAHL, Embase, MEDLINE, ProQuest, PsychInfo, Scopus as well as OpenGrey from onset until 30 April 2022 which will be updated until manuscript submission. The search strategy was designed using medical subject headings (MeSH) as well as Boolean operators “OR”, “AND” or “NOT” appropriately. Customized search strategies were performed for each database. We will also search in trial registers. In the case that a relevant conference abstract is identified, we will contact the authors to obtain full-text article. Reference lists of included studies will be reviewed to identify any relevant additional studies.

#### Search strategy

A search strategy designed with comprehension was used to locate the relevant studies published between January 2000 and April 2022. The search was performed by an experienced information specialist. For piloting purposes, a search was conducted in Medline, and precision tested several times with an aim of yielding the lowest proportion possible of irrelevant studies. The resulting search string was applied to syntaxes of the rest of the data bases where a search was performed. A search for Medline is provided as an example and will be available in the online supplemental appendix 1.

The search terms used were;

1. Malaria or Malaria, Vivax or Malaria, Falciparum or Blackwater Fever or Malaria, Avian
2. Plasmodium or Plasmodium ovale or Plasmodium falciparum or Plasmodium vivax or Plasmodium malariae
3. mosquito
4. culicidae or anopheles or plasmodium or malaria or mosquito or marsh fever or black water fever or paludism
5. 1 OR 2 OR 3 OR 4
6. control, mosquito
7. control, malaria
8. IRS or indoor residual spray or indoor residual spraying
9. (6 OR 7) AND 8
10. 5 AND 9

The search strategy was as well built by an experienced information specialist. We used 15 potentially relevant test articles to test and build the search. The 15 articles were a priori identified using the function similar articles in PubMed and by reading references of the selected articles.

## Data records and management

Following the search, study references will be exported into EndNote referencing software. A review with the name “IRS withdrawal” will be set up in COVIDENCE software, an application for managing systematic literature reviews (https://app.covidence.org/reviews/active). In covidence, the EndNote references’ file will be uploaded. Following the upload, covidence will automatically identify the duplicates and resolve them. Two independent reviewers will screen the titles/ abstracts and then at the full text stage for all the identified records. In case of conflicts/ discrepancies at either stage, they will be resolved through discussions in presence of a third reviewer. During screening, reviewers will be guided by the eligibility criteria (Population, Intervention, Comparators, Outcomes, Publication year, Study design; PICOTS). At the full text level, reviewers will record reasons for exclusion for all excluded studies.

Included studies at both title and abstract as well as at full text will undergo extraction by two independent reviewers after which a consensus will be reached by a member of the technical team of the review. Information to be extracted from the included studies will include study characteristics such as the study title, first author, year of publication, covidence ID, study objective, DOI, abstract, journal name and URL. With regard to geographical information, reviewers will extract the years of sustained IRS use, years when IRS was withdrawn, any other information on IRS use in the area, continent name, country name, country income level, area’s malaria classification (burden category), region name, district name and location name. With regard to methodological information, reviewers will extract data related to the study design, type of effect measure/ measure of association used, method of analysis done, unit of analysis, overall sample size and any assumptions made for the analysis. Study participant characteristics to be extracted will include the age group under study (i.e., children under five years of age, pregnant women, general population, school age children), gender and any other relevant characteristics. Outcomes to be extracted will include malaria incidence and malaria vector abundance. To measure malaria incidence, incidence rates, relative risks, entomological inoculation rates (EIRs), slide positivity rates, test positivity rates may be extracted depending on what the studies will report. Vector abundance on the other hand will be represented by sporozoite rates or mosquito mortality rates. For studies that assess the long-time residual efficacy of insecticides, respective malaria burden indicators both during the implementation phase and after a defined period following stoppage of the intervention will be extracted. A pre-defined data extraction template will be designed, piloted and published in covidence to capture all the study aspects.

### Effect measures

We will report the withdraw effects between the baseline and endline measures depending on the study reported measures. The effects may be reported as a rate ratio or a risk ratio (RR) and accompanying 95% confidence intervals.

### Risk of bias assessment

Appropriate tools will be used to assess the risk of bias for the included studies. Depending on the evidence found, the Cochrane risk of bias 2 [21] and risk of bias in non-randomised studies of interventions tools will be used to assess the risk of bias in the studies measuring the impact of withdrawal. Characteristics will be assessed with respect to sampling methods, the sample size, representativeness of the sample, reliability of the outcomes, analysis methodologies, levels of precision of the indicator measures among others. Risk of bias assessment as well will be done by the two independent reviewers. For both tools, the domains have a signaling question aiming to elicit relevant information. Responses to these questions will be scored for each domain either low risk of bias, some concerns or high risk of bias. The scores of each domain will then be mapped into overall risk-of-bias judgement including categories of low risk of bias, some concerns and high risk of bias.

### Data synthesis and reporting

A meta-analysis will be performed, if sufficient homogeneity exists [22]. Otherwise, results arising from data extraction and risk of bias assessment will be presented using tables and also in form of a narrative synthesis. The results section will among others include a PRISMA flow diagram, table with summarized study characteristics, table of risk of bias assessment as well as findings of the study effect measures. Considering the expected heterogeneity, the results will be pooled using a random-effects model. We will try to minimize the heterogeneity by grouping the studies by setting based on age, the level of endemicity and transmission intensities. Investigation of remaining heterogeneity within a pooled group of studies using a combination of visual inspection of the forest plot along with consideration of the chi-square test (with statistical significance set at p<0.10), and the Higgin’s statistic results according to the recommendations from the Cochrane Handbook. We will perform a sensitivity analysis according to overall study quality i.e., low risk of bias, some concerns and high risk of bias, by comparing random and fixed-effect model and by excluding possible outlier studies, if the visual inspection of the forest plot shows poorly overlapping confidence intervals. Regarding publication bias, funnel plots will be constructed and analysis done using the Egger’s test for analyses that contain more than 10 studies.

### Ethics and dissemination

Since all the studies that we hope to include in this review have probably been published or have got ethical clearance, the nature of this study does not require and has been exempted from Institutional Review Board (IRB) approval. The current review will be published in a peer-reviewed journal and may be used by the National Malaria Control Program to rethink coverage of IRS in malaria endemic areas.

### Patient and public involvement

Patients or the public were not involved in the design, or conduct, or reporting, or dissemination plans of our research.

## Discussion

Malaria remains a disease of public health importance, contributing the largest percentage of inpatient admissions, morbidities and mortalities to the overall population estimates [2]. Indoor IRS forms one of the measures currently being implemented in Africa to control malaria along with long-lasting insecticide treated nets (LLINs) and use of artemisinin-based combination therapies (ACTs) for managing malaria cases. Despite recognition of the benefits/ gains of IRS in the places where it’s well implemented, decisions to withdraw IRS are most of the time not based on evidence such as that from published studies. Several reports have instead indicated that the IRS program particularly in Uganda is largely limited by cost consequences limiting it to only cover about 10% or less of the total population [12]. Given that such withdrawal decisions are most of the time based entirely on cost consequences, information regarding the impact of IRS withdrawal may not be reported. It is also unknown how IRS can be safely withdrawn without causing an upsurge/ outbreak of malaria in areas where use of the intervention has previously been sustained. As we look towards achieving a “zero malaria” world from 2021-2025, there is need to continue scaling up malaria control interventions some of which may include increase in coverage for IRS programs. Collecting primary data for IRS may be tense, time consuming and expensive since the intervention is implemented on a rather large scale and because the withdrawal may not be pre-planned hence taking on a kind of naturally occurring event. For this reason, it is also anticipated that studies may rely heavily on administrative data to assess the impact of IRS withdrawal. Given that such administrative data may be health facility data, we can rely on such data since it provides real world evidence that can be based on to inform programs such as IRS. The current review will considerably contribute to the evidence base since it will provide data that will be a result of systematic and explicit methods that identify, select and critically appraise research.

This systematic review will help update the knowledge on the impact of IRS withdrawal and the need to design a clear exit strategy. In addition, we will use rigorous methodology in accordance with the Cochrane handbook and the results will be reported as stated by the PRISMA statement. Therefore, we will provide relevant knowledge that will inform the relevancy of IRS exit strategies as the countries move towards malaria elimination. However, the certainty of the evidence of this systematic review may be limited by the limited number of studies available and the possible low quality of the individual studies.

## Data Availability

This being a systematic review protocol, no data have been produced.

## Declarations

### Availability of Data and Materials

Not applicable (this is a protocol for a systematic review)

## Funding

None

## Competing Interests

We declare that no competing interests exist.

## Authors’ Contribution

PO, EK, IK, PL, PK, PK conceptualized the idea. PO wrote the initial protocol, PO, EK and CN developed the search strategy. EK, PL, IK and RM have reviewed and approved the protocol for publication. All authors read and approved the final version of the protocol.

## Acknowledgement

We would like to acknowledge and thank the ACRES team for their unwavering support throughout the entire process of writing this protocol.

## Appendix I

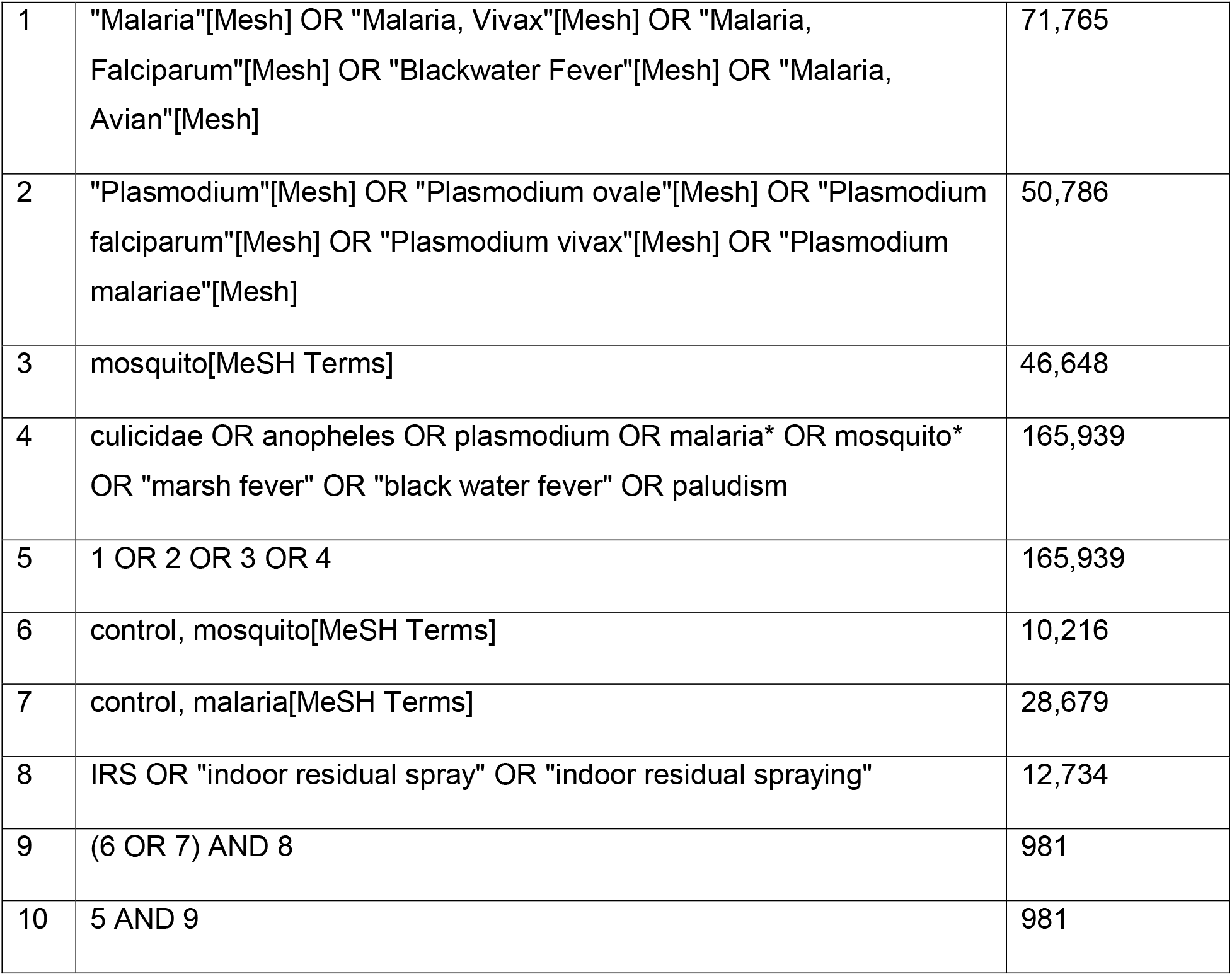
Medline.

## Notes

### Competing Interest Statement

The authors have declared no competing interest.

### Funding Statement

The study did not receive any funding

