## Supplemental Table 1 for "Vulnerability of Populations to Malaria after Indoor Residual Spraying is Withdrawn from Areas where its Use has Previously Been Sustained. Protocol for a Systematic Review"

Medline search strategy

| | | |
|---|---|---|
| 1 | "Malaria"[Mesh] OR "Malaria, Vivax"[Mesh] OR "Malaria, Falciparum"[Mesh] OR "Blackwater Fever"[Mesh] OR "Malaria, Avian"[Mesh] | 71,765 |
| 2 | "Plasmodium"[Mesh] OR "Plasmodium ovale"[Mesh] OR "Plasmodium falciparum"[Mesh] OR "Plasmodium vivax"[Mesh] OR "Plasmodium malariae"[Mesh] | 50,786 |
| 3 | mosquito[MeSH Terms] | 46,648 |
| 4 | culicidae OR anopheles OR plasmodium OR malaria* OR mosquito* OR "marsh fever" OR "black water fever" OR paludism | 165,939 |
| 5 | 1 OR 2 OR 3 OR 4 | 165,939 |
| 6 | control, mosquito[MeSH Terms] | 10,216 |
| 7 | control, malaria[MeSH Terms] | 28,679 |
| 8 | IRS OR "indoor residual spray" OR "indoor residual spraying" | 12,734 |
| 9 | (6 OR 7) AND 8 | 981 |
| 10 | 5 AND 9 | 981 |
